# Gentrification and Food Environments: A Rapid Evidence Assessment

**DOI:** 10.1101/2023.03.07.23286919

**Authors:** Simone Gie, Fiona Borthwick

## Abstract

Gentrification is a complex and controversial process, where the influx of new, wealthier residents to previously run-down neighbourhoods brings change such as economic development, infrastructure investments and lower crime rates, but can be to the detriment of the original lower-income residents, who are either displaced, or stay but cannot take advantage of the new opportunities. Understanding how neighbourhood change affects food environments can shed light on the possible causal pathways between gentrification and urban health inequalities. This rapid evidence assessment reviewed evidence on the impact of gentrification on the healthfulness of food environments globally. Ten studies were identified through a systematic keyword search and assessed. We found limited evidence of an effect, with a small, albeit consistent, body of evidence mostly comprised of low- to medium-quality observational studies, all from high-income countries. Most studies examined effects on availability or affordability of food, finding an association between gentrification and increased availability of unhealthy foods, or reduced affordability for original low-income residents.

## 1 Introduction

Gentrification is a complex and controversial process, where the influx of new, wealthier residents to previously run-down neighbourhoods brings change such as economic development, infrastructure investments and lower crime rates, but can be to the detriment of the original lower-income residents, who are either displaced, or stay but cannot take advantage of the new opportunities (Rhodes-Bratton *et al*. 2018).

Food environments are defined by Franco et al. (2016) as all aspects of the local environment that influence dietary behaviours. Neighbourhood changes that occur with gentrification, such as the replacement of local “mom and pop” stores with upmarket boutiques and retail chains (Krase and DeSena 2020), therefore may also influence characteristics of the food environment. As dietary intake is a key determinant of diet quality, nutrition status and disease (Afshin *et al*. 2019), understanding how food environments are affected by gentrification can shed light on the possible causal pathways between gentrification and health inequalities.

This rapid evidence assessment (REA) aims to review the evidence on the impact of gentrification on food environments and was conducted according to guidance from the UK government’s Department of Environment Food and Rural Affairs (DEFRA) (Collins *et al*. 2015) and the Guideline for Rapid Evidence Assessments in Management and Organizations by the Centre for Evidence-Based Management (CEBMa) (Barends *et al*. 2017).

To our knowledge, this is the first review of the impact of gentrification on food environments. Previous reviews have focused on measures of the food environment (Lytle and Sokol 2017); nutrition interventions in low-income rural and urban retail environments (Fergus et al. 2021); community-level interventions to improve access to nutritious food in low and middle income countries (LMICs) (Durao et al. 2020); socioeconomic differences in the association between the food environment and diet (Mackenbach et al. 2019); mapping evidence from projects on drivers of food choice to a food environment framework (Constantinides *et al*. 2021); associations between food environment characteristics and diet, nutrition and health outcomes in urban LMIC settings (Westbury *et al*. 2021); and the state of food environment research in LMICs (Turner *et al*. 2020).

We start by defining a clear research question, then describe the methodology used to identify and evaluate the literature, provide a judgement on the quality of evidence, and summarize salient themes. We conclude by highlighting gaps in the literature for future research consideration.

## 2 Definitions

While there are competing definitions for *gentrification* (Tulier *et al*. 2019), the term is generally understood as the process in which a poor area experiences an influx of high-income newcomers who drive up property values, often resulting the displacement of original, low-income residents (Merriam-Webster 2020).

Related concepts of gentrification include *tourism gentrification*, where neighbourhoods change to suit the needs of wealthy visitors (Loda *et al*. 2020, Sánchez-Ledesma *et al*. 2020); *commercial gentrification* where retail change occurs but is disconnected from residential gentrification (Kosta 2019); and *ecological gentrification*, the pursuit of an environmental agenda related to public green spaces that leads to the displacement of homeless people (Dooling 2009).

Often used interchangeably with gentrification (Tulier *et al*. 2019), *urban renewal* refers to programmes to restore degraded buildings (Merriam-Webster 2020), which frequently displaces original residents and leads to gentrification (Komakech and Jackson 2016).

The *food environment* is defined Swinburn *et al*. (2013) as the “collective physical, economic, policy and sociocultural surroundings, opportunities and conditions that influence people’s food and beverage choices and nutritional status.”

Other recent work has expanded on past definitions of food environments to encompass the reality in LMICs. Turner *et al*. (2018) describe food environments as the “interface where people interact with the wider food system to acquire and consume foods”. This conceptualization includes both market and non-market food sources and splits food environments into external (e.g. availability, price) and personal (e.g. accessibility, affordability) domains.

Downs *et al*. (2020) propose a definition applicable to both LMICs and high-income settings: “The consumer interface with the food system that encompasses the availability, affordability, convenience, promotion and quality, and sustainability of foods and beverages in wild, cultivated, and built spaces that are influenced by the socio-cultural and political environment and ecosystems within which they are embedded.” The incorporation of different food system typologies (natural and built) aims to better reflect the reality of how people interface with food systems in diverse settings.

The High Level Panel of Experts on Food Security and Nutrition (HLPE 2017) outlines four domains of the food environment, which have been used as a framework in this REA:

- Availability and physical access (proximity)
- Affordability (both absolute prices and relative to purchasing power)
- Promotion, advertising and information
- Food quality and safety (This dimension is expanded on and described by Herforth and Ahmed (2015) as ‘desirability’, and Caspi *et al*. (2012) as ‘acceptability’)

Affordability is measured in food environment studies either as absolute (e.g. food prices) or relative to income and purchasing power (Lee *et al*. 2013, Herforth and Ahmed 2015, Franco *et al*. 2016). By this logic, where household income is the denominator of affordability, foods of the same price can have different affordability for different households in the same neighbourhood. The affordability of food environments is therefore subjective, and factors impacting household income can affect the affordability of food even when prices remain static. This idea is expressed in Turner *et al*. (2018)’s conceptualization of food environments where price is a dimension of the external food environment while affordability is a dimension of the personal food environment.

*Food mirages* refer to areas where food outlets are plentiful but unaffordable for low-income residents (Breyer and Voss-Andreae 2013).

The terms *healthy* and *unhealthy* to describe food outlets or environments in this REA follow the authors’ categorization. ‘Unhealthy’ usually defines neighbourhoods with high concentrations of fast food and convenience stores, also known as ‘food swamps’, where areas are overwhelmed with opportunities to access high calorie food and beverages, (Bridle-Fitzpatrick 2015), and/or neighbourhoods lacking access to healthy foods such as fruit and vegetables, also known as ‘food deserts (Widener and Shannon 2014). *Food outlets* is used to describe all food acquisition opportunities (retail and catering).

## 3 Methodology

### 3.1 Research question

The aim of this REA is to review what is known about the link between gentrification and food environments, specifically asking *How does gentrification impact the healthfulness of food environments?* The lens of food environments as experienced by original or low-income residents was applied to the research question.

The method applied by this REA was based on guidance from DEFRA set out by Collins et al (2015), and supplemented by guidance from CEBMa (Barends *et al*. 2017). Guidance on applying the SPICE framework was taken from Booth (2006) and Wilson *et al*. (2016). This assessment can be viewed as a streamlined REA report, rather than a full scoping report.

Applying a streamlined approach to identifying and assessing recent peer reviewed evidence while making recommendations for further systematic reviews allows for a quicker appraisal of a question and can ascertain the potential structure of a full systematic review. This approach to REAs offers researchers and policymakers a robust additional method to identifying evidence around a topic within a timeframe of weeks, and so can help to respond to rapidly emerging issues and help to define the terms of scoping reports as well as strategic evidence assessments.

Application of Booth (2006)’s SPICE framework for defining research questions (Table 1) allowed elaboration of a clear research question and search terms.

**Table 1:** Application of the SPICE framework to review the association between gentrification and healthy food environments

| SPICE element | Relevant search terms or inclusion/exclusion concept | Justification |
| --- | --- | --- |
| **Setting** | Urban areas<br>All countries | Gentrification literature is geographically biased towards North America and Western Europe, but is a global phenomenon (Krase and DeSena 2020) |
| **Population** | Urban residents<br>All socio-economic groups | Original low-income residents who remain after gentrification were of particular interest |
| **Intervention** | Gentrification<br>Urban renewal<br>Urban regeneration | These terms are often used interchangeably in the literature (Tulier *et al.* 2019)<br>Urban renewal can lead to gentrification (Komakech and Jackson 2016) |
| **Comparator** | Before gentrification<br>Similar non-gentrified neighbourhoods | Interested in the effect of gentrification compared to absence of gentrification |
| **Evaluation** | Healthy food environments<br>Presence of fast-food outlets<br>Presence of supermarkets<br>Ratio of healthy/unhealthy foods available in retail outlets | Health, nutrition and dietary intake outcomes (which may be impacted through other non-food pathways e.g. green spaces for exercise, access to health services) were not of interest and were excluded in order to isolate the effect on food environments |

### 3.2 Study selection

Figure 1 outlines the study selection process. Search terms and concepts identified in the SPICE framework were used to develop a search string combined with Boolean operators ‘AND’ and ‘OR’, and to inform inclusion and exclusion criteria. The streamlined approach applied in this case reduced the time limit for publication from 10 to 5 years, focused on the 100 most relevant articles in the search, and selected articles only in English. The justification for a streamlined approach is to allow for a faster identification of a highly selected range of evidence which can then inform recommendations for full systematic reviews. The limitations of these restrictions are discussed.

**Figure 1:**
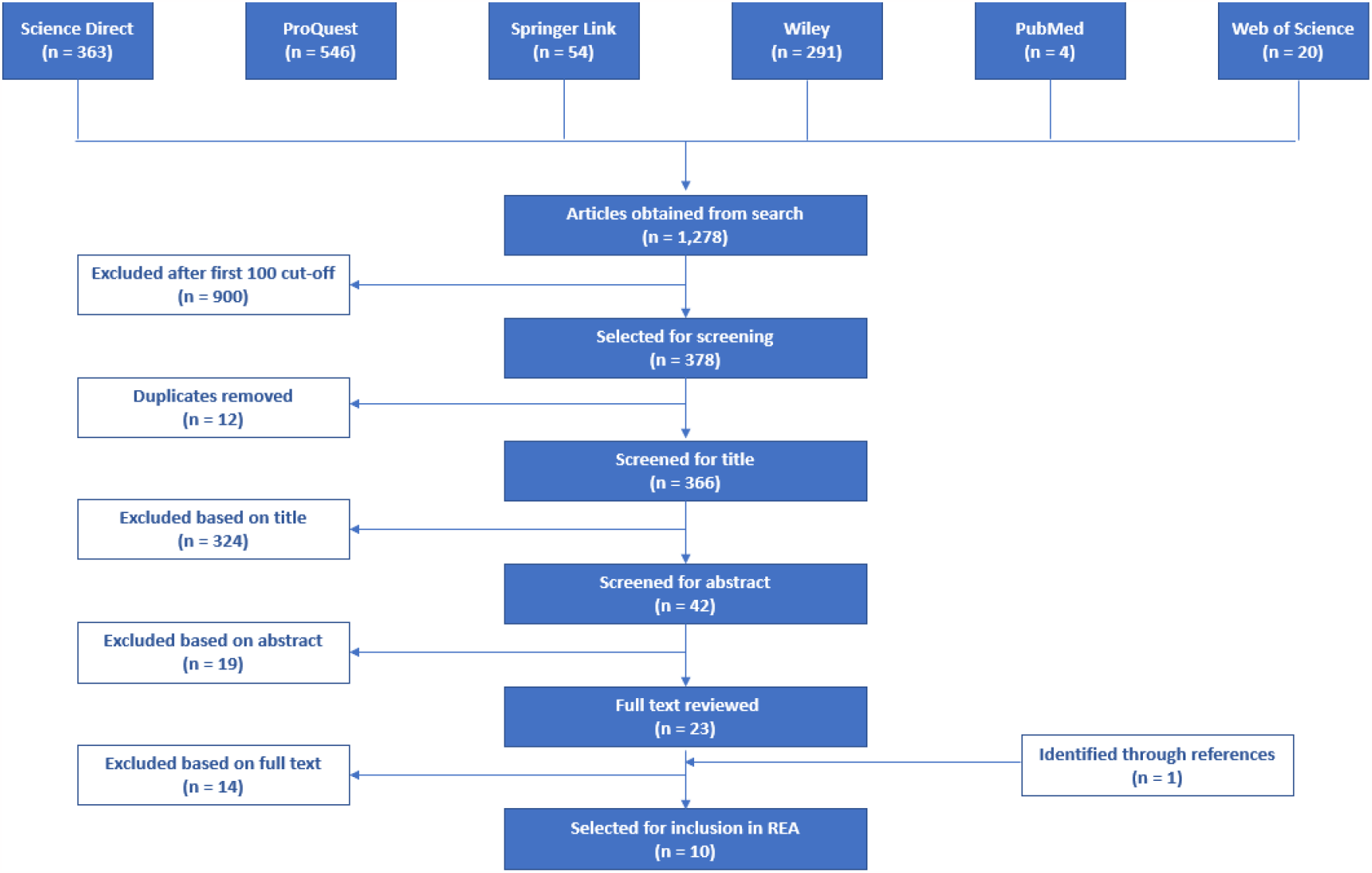
Study selection process

The search string entered into databases was:

*(gentrification OR “urban renewal” OR “urban regeneration” OR “neighbourhood renewal”) AND (“food environment” OR retail OR “fast food” OR supermarket) AND food*

The search string was applied to six databases (shown in Figure 1) in September 2020, according to the REA method employed, described above. Filters for articles published in the last five years, research articles/journals only, full text, peer reviewed and English language were applied where available, generating a total of 1,278 results.

After sorting for relevance, the first 100 articles from the three databases yielding over 100 results, and all articles from databases yielding fewer than 100, were imported into EndNote X9 (n=378).

After removing duplicates (n=12), 366 titles were screened and 42 were retained. Abstracts were reviewed against inclusion and exclusion criteria (Table 2) and 23 articles were retained, plus an additional article (from 2013) was identified via reviewing reference lists of all selected articles and included for relevance. After reading the full studies, ten were selected which met the scope of the REA.

**Table 2:**
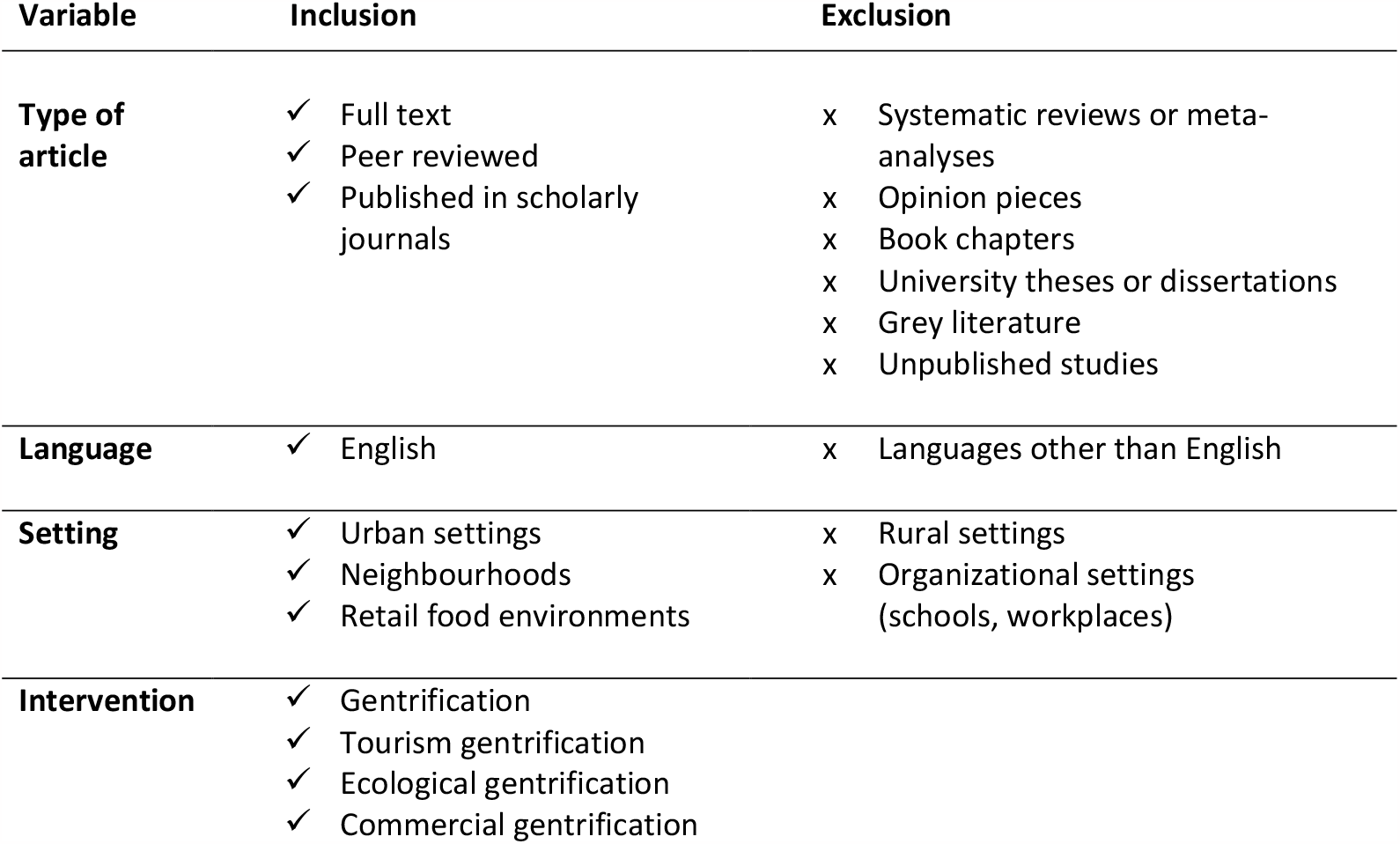
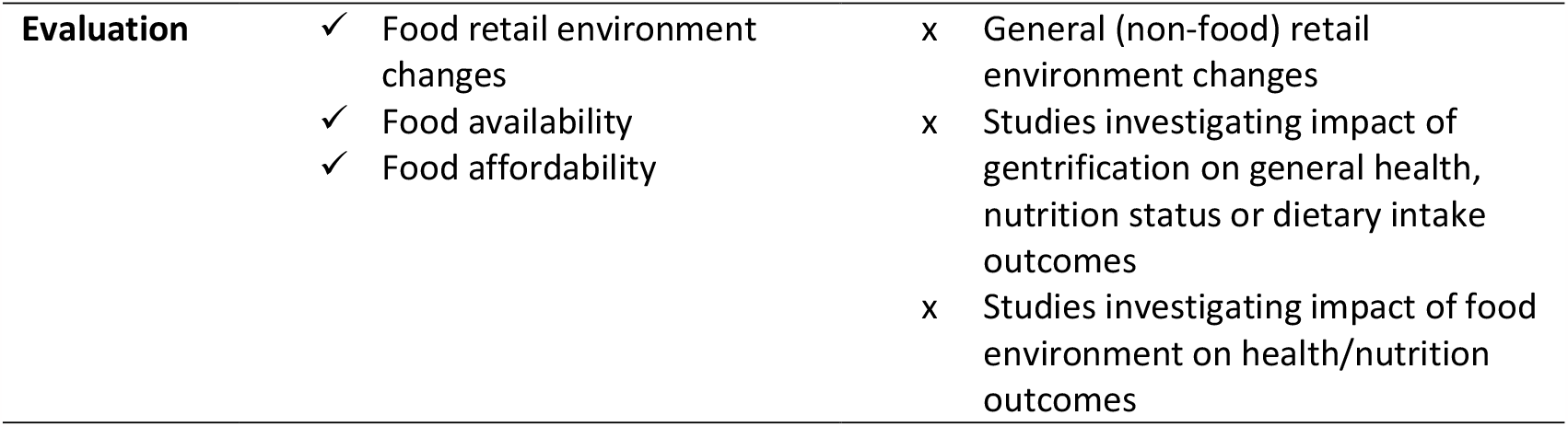
Study inclusion and exclusion criteria

| Variable | Inclusion | Exclusion |
| --- | --- | --- |
| **Type of article** | ✓ Full text<br>✓ Peer reviewed<br>✓ Published in scholarly journals | x Systematic reviews or meta-analyses<br>x Opinion pieces<br>x Book chapters<br>x University theses or dissertations<br>x Grey literature<br>x Unpublished studies |
| **Language** | ✓ English | x Languages other than English |
| **Setting** | ✓ Urban settings<br>✓ Neighbourhoods<br>✓ Retail food environments | x Rural settings<br>x Organizational settings (schools, workplaces) |
| **Intervention** | ✓ Gentrification<br>✓ Tourism gentrification<br>✓ Ecological gentrification<br>✓ Commercial gentrification |  |

**Table 2:** Study inclusion and exclusion criteria
|  |  |  |  |  |
| --- | --- | --- | --- | --- |
| Evaluation | ✓ | Food retail environment changes | x | General (non-food) retail environment changes |
|  | ✓ | Food availability | x | Studies investigating impact of gentrification on general health, nutrition status or dietary intake outcomes |
|  | ✓ | Food affordability | x | Studies investigating impact of food environment on health/nutrition outcomes |

### 3.3 Study review

One author reviewed the ten studies and extracted details on methods, findings and key themes. As randomized trials are difficult and rare in neighbourhood food environment studies (Lytle 2009), the UK Government’s ‘How To Note: Assessing the strength of evidence’ (DFID 2014), referred to hereon as *the How To Note*, was considered an appropriate tool for evidence evaluation. The *How To Note* provides a robust framework for evaluating evidence generated by all research designs, including experimental, observational, quantitative and qualitative studies. The process used in a DFID REA (Cramer *et al*. 2016) was used as a template, described as follows.

A checklist was adapted from the *How To Note’s* checklist of quality assessment. Many concessions must be made in order for an REA to be conducted rapidly (Barends *et al*. 2017). In order to adapt to the time and personnel constraints of this REA, two principles (reliability and cultural sensitivity, referring to research designs that fail to consider local, cultural factors that might affect behaviours and trends) were removed from assessment.

Following the DFID example REA, a grading system was devised to ensure a structured approach. Using checklist questions as a guide, two reviewers independently assessed the ten articles, giving a grade of 1 to 3 for each principle (1 being major concerns to 3 being no concerns). Each study was then assigned an average score assuming equal weighting for each principle, and categorized as low (<2.0), medium (2.0-2.5), or high (>2.5) quality, with cut-offs decided by the reviewer. A narrative approach was used to synthesize the findings.

## 4 Results

### 4.1 Summary of studies

Selected studies are detailed in Table 3. Of the ten studies, seven (Breyer and Voss-Andreae 2013, Anguelovski 2015, Whittle *et al*. 2015, Komakech and Jackson 2016, Rhodes-Bratton *et al*. 2018, Berger *et al*. 2019, Kosta 2019) were conducted in North America and three (Bilal *et al*. 2018, Loda *et al*. 2020, Sánchez-Ledesma *et al*. 2020) in Western Europe.

**Table 3:**
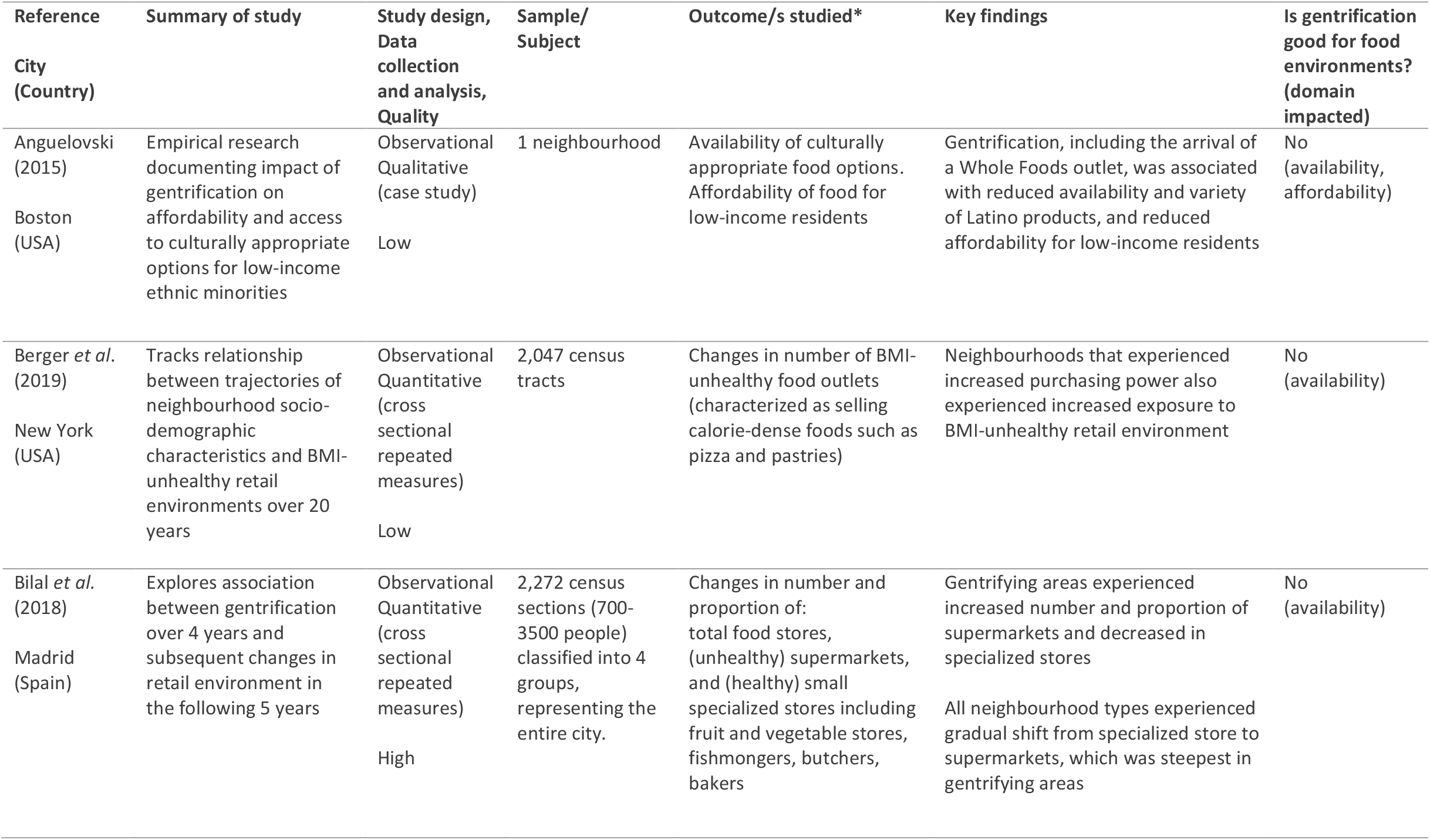
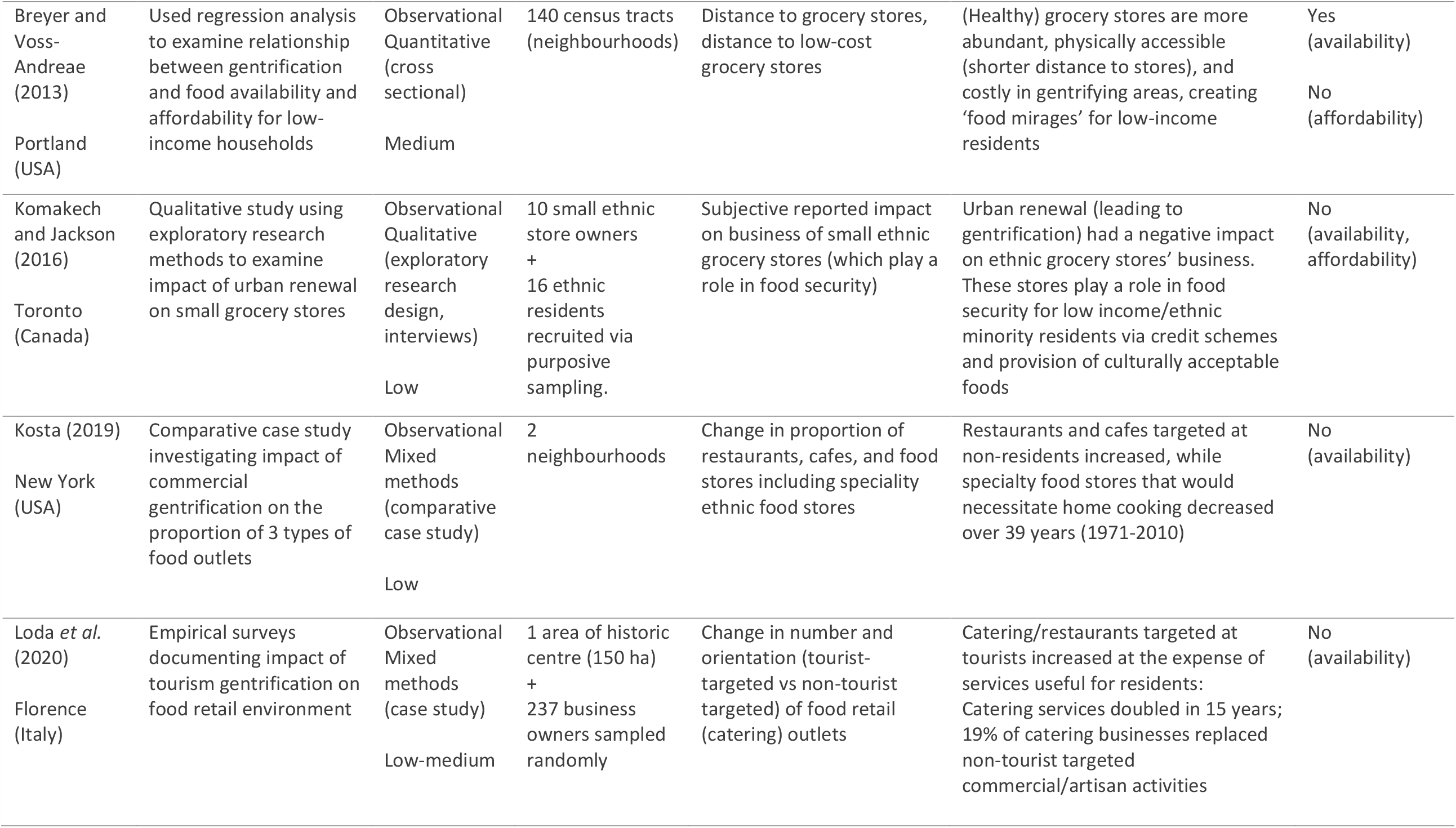
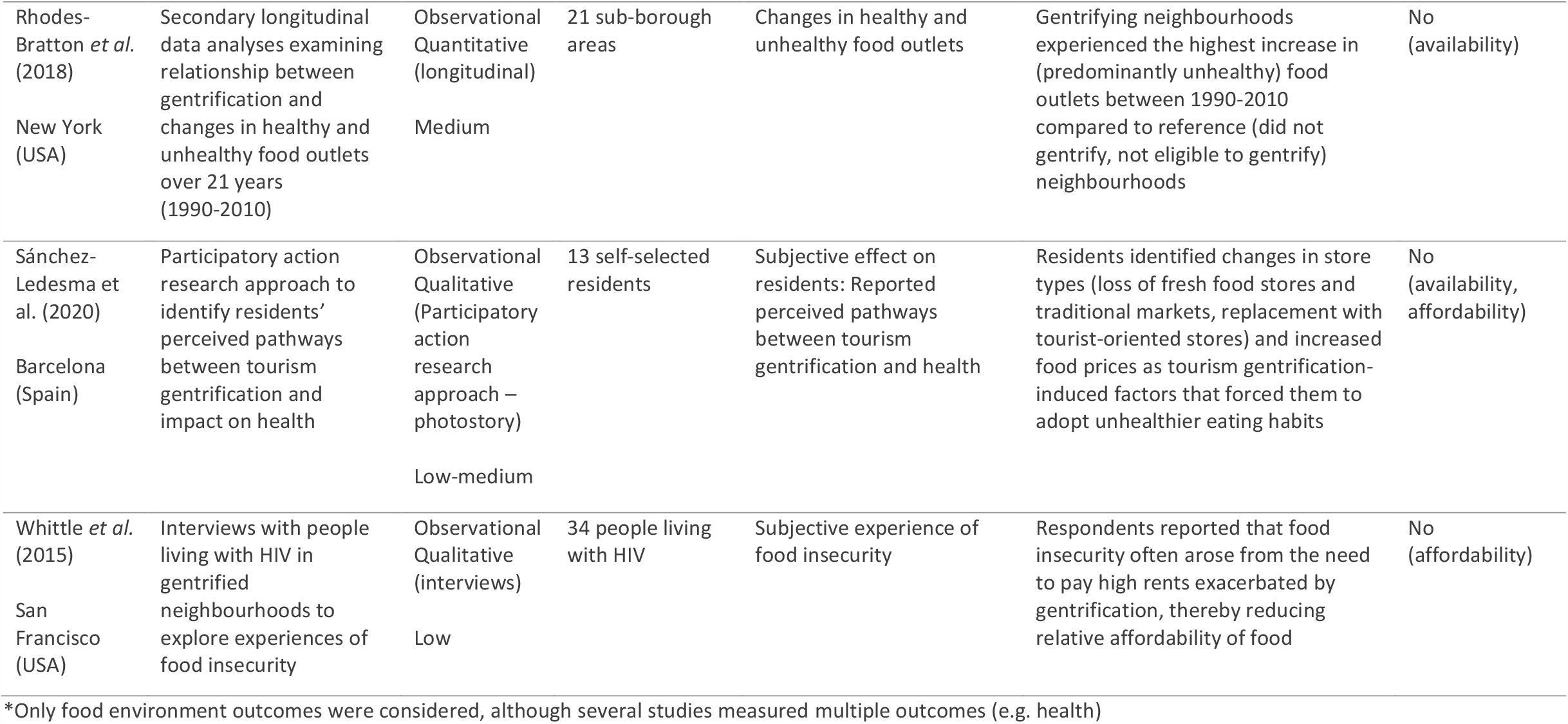
Summary of studies

All studies used observational research designs, four (Anguelovski 2015, Whittle *et al*. 2015, Komakech and Jackson 2016, Sánchez-Ledesma *et al*. 2020) used qualitative data in their analysis, four (Breyer and Voss-Andreae 2013, Bilal *et al*. 2018, Rhodes-Bratton *et al*. 2018, Berger *et al*. 2019) used quantitative, and two (Kosta 2019, Loda *et al*. 2020) used mixed methods.

Six studies (Breyer and Voss-Andreae 2013, Anguelovski 2015, Bilal *et al*. 2018, Rhodes-Bratton *et al*. 2018, Berger *et al*. 2019, Kosta 2019) used a neighbourhood, census tract or other geographical boundary as the unit of analysis, three qualitative studies (Whittle *et al*. 2015, Komakech and Jackson 2016, Sánchez-Ledesma *et al*. 2020) used residents as subjects, and one study (Loda *et al*. 2020) used both.

Nine studies (Breyer and Voss-Andreae 2013, Anguelovski 2015, Komakech and Jackson 2016, Bilal *et al*. 2018, Rhodes-Bratton *et al*. 2018, Berger *et al*. 2019, Kosta 2019, Loda *et al*. 2020, Sánchez-Ledesma *et al*. 2020) explored the effect of gentrification (or socioeconomic status (SES) as a proxy), on one or more domains of the food environment. The most common outcome measured, in five studies, (Rhodes *et al*. 2009, Bilal *et al*. 2018, Berger *et al*. 2019, Kosta 2019, Loda *et al*. 2020) was change in types of food outlets using repeated cross-sectional measures or longitudinal data. The tenth study (Whittle *et al*. 2015) began with the outcome, investigating food insecurity and identifying gentrification as a driver of reduced affordability of foods.

All studies concluded that gentrification had a negative effect on at least one domain of the food environment when considering the subjective experience of original or low-income residents.

## 5 Evaluation of Evidence

### 5.1 Evaluation of individual studies

Table 4 displays the results of the quality checklist.

**Table 4:**
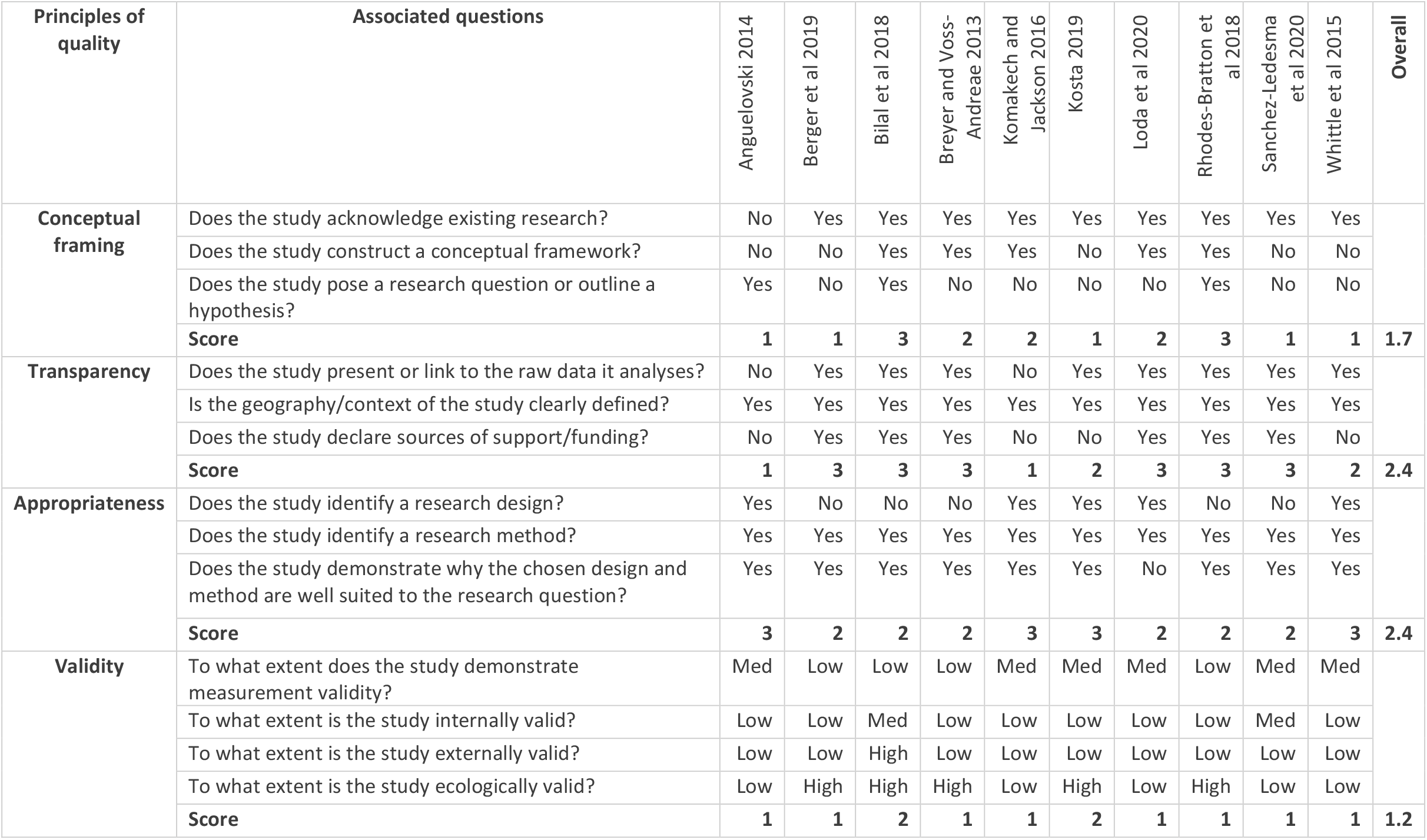
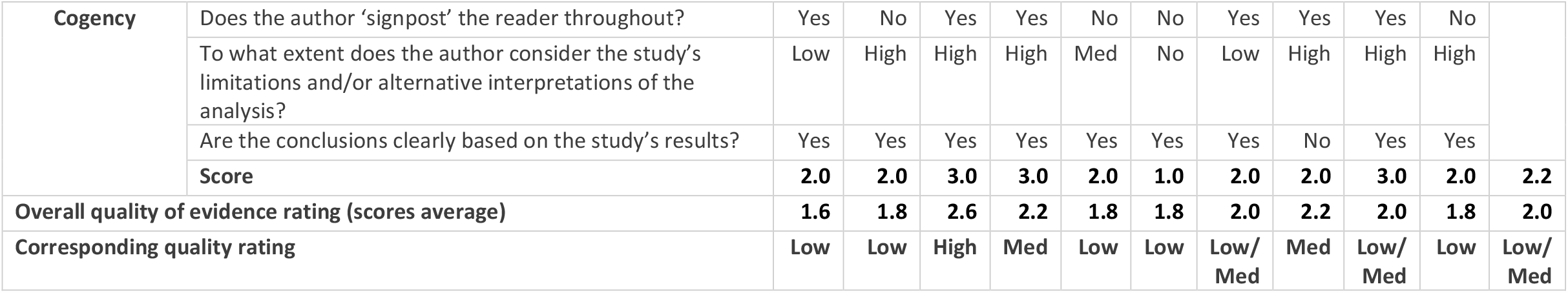
Quality of evidence checklist for studies, adapted from DFID (2014) **Scores:** 3 = no concerns 2 = some concerns 1 = major concerns **Quality cut-offs for averages:** <2.0 = low 2.0-2.5 = medium >2.5 = high

On average, the sample was judged to be low quality for conceptual framing, with only two studies (Bilal *et al*. 2018, Rhodes-Bratton *et al*. 2018) considered to fully meet all three criteria of acknowledging existing research, constructing a conceptual framework and posing a research question or outline a hypothesis.

The sample was judged to be medium quality for transparency, with six studies (Breyer and Voss-Andreae 2013, Bilal *et al*. 2018, Rhodes-Bratton *et al*. 2018, Berger *et al*. 2019, Loda *et al*. 2020, Sánchez-Ledesma *et al*. 2020) judged to fully meet all three criteria of presenting or linking to the raw data, clearly defining the geography or context of the study and declaring sources of funding.

The body of evidence was evaluated as medium quality for appropriateness, with four studies (Anguelovski 2015, Whittle *et al*. 2015, Komakech and Jackson 2016, Kosta 2019) judged to meet all criteria of identifying a research design and method and demonstrating why the chosen design and method was well suited to the research question.

Studies were considered to perform poorly for validity, with only two studies (Bilal *et al*. 2018, Kosta 2019) considered to demonstrate all considered forms of validity (measurement, internal, external and ecological).

The sample was evaluated to be medium quality for cogency, with three studies (Breyer and Voss-Andreae 2013, Bilal *et al*. 2018, Sánchez-Ledesma *et al*. 2020) graded highly for signposting the reader, considering the study’s limitations or alternative interpretations of the analysis, and basing conclusions clearly on the study’s results.

### 5.2 Evaluation of the body of evidence

Using the DFID *How To Note*, the overall body of evidence was judged on quality, size, context and consistency.

The quality assessment described above judged five studies (Anguelovski 2015, Whittle *et al*. 2015, Komakech and Jackson 2016, Berger *et al*. 2019, Kosta 2019) to be low quality, two (Loda *et al*. 2020, Sánchez-Ledesma *et al*. 2020) to be low-medium, two (Breyer and Voss-Andreae 2013, Rhodes-Bratton *et al*. 2018) to be medium, and one (Bilal *et al*. 2018) to be high quality. The overall quality of the sample was therefore judged to be low-medium. However the range of different designs used, which triangulates findings, is a strength (DFID 2014).

Although REAs do not involve a comprehensive review of the literature, summarizing the characteristics of the body of evidence include some subjective judgement of the size of the body of evidence (DFID 2014). The size of the evidence base was considered be small, with only ten studies identified. Although there are no specific numbers that constitute size DFID (2014), a crude test on ScienceDirect comparing results elicited from the search terms *gentrification “food environment”* (n=34) and *gentrification “mental health”* (n=382) provides a basic indication of the relative size of the evidence body compared to other gentrification-related topics.

The body of evidence is context-specific (as opposed to global), heavily skewed towards North America then Western Europe, and totalling just four countries (USA, Canada, Spain and Italy). A convincing body of evidence would ideally exist globally as well as in the context of interest. Without a comparison group in different settings, context-related factors may confound findings (DFID 2014). The absence of studies from LMICs, despite the global focus of the search, was surprising given the nutrition transition, urbanization and gentrification occurring in these regions, and that food environment research is gaining traction in LMICs (Turner *et al*. 2020). This may have been due to the limitations of the search imposed by the REA methodology.

The body of evidence, however, was consistent, with all studies concluding that gentrification had a negative effect on food environments, particularly availability and affordability, when considered through the lens of low-income groups. However, this could suggest publication bias, where studies reporting a significant relationship are more likely to be published than those with null results (Caspi et al. 2012).

#### Measurement validity

Five studies (Breyer and Voss-Andreae 2013, Bilal *et al*. 2018, Rhodes-Bratton *et al*. 2018, Berger *et al*. 2019, Kosta 2019) relied exclusively on secondary data (e.g. business directories or geographic information system-based methods) to characterise the food environment, which Liese et al. (2013) found results in significant error. Both Liese *et al*. (2013) and Kosta (2019) recommend combining these data with field census or other methods such as qualitative interviews, however this was only done in one study (Loda et al. 2020).

The classification of outlets as ‘healthy’ or ‘unhealthy’ was measured at the level of store type. More precise retail-level data (such as measures of relative shelf space, availability and affordability of specific foods, etc.) are likely required.

Gentrification, which is complex, non-linear and phased, is also inherently problematic to study (Tulier et al. 2019), and was measured inconsistently across studies, with some, e.g. Rhodes-Bratton *et al*. (2018), relying on secondary gentrification rankings others, e.g. Berger *et al*. (2019), using sociodemographic data such as change in Black and Hispanic populations.

#### Internal validity

Three studies (Breyer and Voss-Andreae 2013, Bilal *et al*. 2018, Berger *et al*. 2019) analysed cross-sectional data (two with repeated measures) which has limited capacity to demonstrate cause and effect (Lytle 2009). Gentrification may impact food environments, but the reverse may also be true, such as when the opening of new supermarkets makes a neighbourhood more attractive to wealthy newcomers (Cohen 2018).

Only one study (Bilal et al. 2018) aimed to control for causal direction by analysing neighbourhood change and subsequent retail change in two separate time periods. Confounding remained an issue, however, as study periods overlapped with recession and recovery (Bilal *et al*. 2018).

Use of longitudinal data or repeated measures of cross-sectional data do not resolve the issue of confounding as neighbourhoods themselves also change over time (Lytle 2009). The four studies using residents (Whittle *et al*. 2015, Komakech and Jackson 2016, Loda *et al*. 2020, Sánchez-Ledesma *et al*. 2020) were also prone to confounding, as people are not ‘randomly assigned’ to neighbourhoods, but may live there due to income, proximity to work, or other factors (Lytle 2009).

Although conceptualization of causal mechanisms is essential to inform policy (Tulier et al. 2019), only three studies identified potential causal pathways: increased property value driving out small retailers (Bilal *et al*. 2018); exodus of ethnic families reducing demand for ethnic retailers (Komakech and Jackson 2016); and high rents reducing purchasing power, the denominator of food affordability, of vulnerable people (Whittle *et al*. 2015).

In summary, this REA has found that the evidence body linking gentrification with unhealthier food environments is small, albeit consistent, and of low to medium quality. This corresponds most closely to DFID’s description of ‘limited evidence’, characterized by mostly medium to low quality observational studies.

## 6 Summary of key themes

Four themes emerged from the studies reviewed: availability, affordability (food mirages), cultural relevance, and catering to a transient population. Breaking down findings into food environment domains helps distinguish which associations are the most robust (Caspi et al. 2012).

### 6.1 Availability of healthy and unhealthy food

Nine studies explored the concept of availability, of which the majority (Bilal et al. 2018, Rhodes-Bratton et al. 2018, Berger et al. 2019, Kosta 2019, Loda et al. 2020) measured changes in the number and/or proportion of healthy versus unhealthy food outlets, and one (Breyer and Voss-Andreae 2013) measured distance to (healthy) grocery stores.

Seven of these nine studies (Anguelovski 2015, Komakech and Jackson 2016, Bilal *et al*. 2018, Berger *et al*. 2019, Kosta 2019, Loda *et al*. 2020, Sánchez-Ledesma *et al*. 2020) found gentrification to be associated with increased availability of unhealthy foods and/or decreased availability of healthy or culturally appropriate foods. One (Rhodes-Bratton *et al*. 2018) found increased availability of both healthy and unhealthy, and one (Breyer and Voss-Andreae 2013) found increased availability of healthy (albeit unaffordable) food.

The categorization of food outlet types as healthy or unhealthy differed by study. Supermarkets were considered unhealthy in Madrid, as they were more likely to offer low-cost processed foods (Bilal *et al*. 2018), but were labelled healthy in the American studies, where they are assumed to carry more healthy options compared to convenience stores (Franco *et al*. 2016).

This differing classification of store type in each context hinders comparability and thus meta-analysis of effect estimates. Categorization of ‘healthy’ and ‘unhealthy’ at the store level could lead to measurement error and inconsistent findings, as stores may offer both healthy and unhealthy options. Caspi *et al*. (2012) argue that since supermarkets offer both fresh and ultra-processed foods, applying this dichotomous classification may be overly simplistic. Consumer-level retail measures such as shelf space and product placement would a provide more granular understanding.

After their systematic review found consistent evidence of an association between availability and dietary behavior in LMICs, which contrasted with previous findings from high-income countries (HICs), Westbury *et al*. (2021) hypothesized that availability may be more important in LMICS than HICs. The authors suggested that this could be due in part to access to transport which makes it easier for people to buy food outside their neighbourhoods. Applying the same consideration to gentrifying neighbourhoods, if poorer residents are less likely to have access to private transport, food availability may be an important predictor of dietary behaviors.

### 6.2 Food mirages: unaffordable abundance

Three low quality studies (Anguelovski 2015, Whittle *et al*. 2015, Komakech and Jackson 2016), one low-medium quality study (Sánchez-Ledesma *et al*. 2020) and one medium quality study (Breyer and Voss-Andreae 2013) explored the issue of affordability for original residents. Three of these five (Breyer and Voss-Andreae 2013, Anguelovski 2015, Komakech and Jackson 2016) considered prices relative to the purchasing power of certain groups. One paper (Whittle *et al*. 2015) explored a mechanism on the demand side, whereby high rents due to gentrification in San Francisco reduced the food budgets of people living with HIV. All studies concluded that food affordability worsened with gentrification for the populations considered.

Unaffordability often coincided with abundant availability, exemplifying the concept of ‘food mirages’, where food outlets are plentiful but unaffordable for low-income residents (Breyer and Voss-Andreae 2013). Breyer and Voss-Andreae (2013) found shorter distances to grocery stores and more abundant but costly food in gentrified areas, pointing out that these areas would not appear problematic from a standard food desert perspective.

Constantinides *et al*. (2021), who found that gender dynamics was an important factor in LMIC food environment studies, argued for applying an equity lens to assessment of the personal food environment. The above findings support this argument and suggest that considering equity may help understand how the personal circumstances of poorer residents, such as income or time available for food preparation, mediate how external food environments in gentrified areas are experienced.

None of the studies reviewed differentiated between the relative affordability of healthy versus unhealthy food, with the partial exception of Sánchez-Ledesma *et al*. (2020) who found that increased food prices led to self-reported ‘worse nutrition habits’ among residents. Since healthy diets have been found to cost more than unhealthy ones (Rao *et al*. 2013), it could therefore be assumed that any issue with affordability of food in general would be exacerbated if only healthy foods were considered. If studies investigating affordability fail to make this distinction, findings may have limited value in explaining obesogenic food environments.

### 6.3 Cultural acceptability of available food

Two low quality studies (Anguelovski 2015, Komakech and Jackson 2016) looked at the theme of cultural acceptability, with both concluding that gentrification led to decreased access to affordable and culturally preferred items for ethnic minorities, such as halal foods, via the closure of stores.

The concept of cultural preferences is largely absent from food environment definitions, aside from Herforth and Ahmed (2015)’s dimension of ‘desirability’ which includes cultural norms. Caspi *et al*. (2012) argue that food environment constructs should be expanded to include cultural relevance, which may be significant in areas with large immigrant populations.

Other aspects of acceptability did not appear in the studies. This aligns with a systematic review of food environment research in LMICs by (Turner *et al*. 2020) which found aspects of the personal food environment such as desirability and convenience to feature less prominently than the external food environment. Caspi *et al*. (2012) also concluded that food acceptability in general is understudied in food environment literature.

### 6.4 Catering for transient populations

Three low-medium and low quality studies looked at specific types of gentrification: tourism gentrification in Florence and Barcelona, where neighbourhoods change to suit the needs of wealthy visitors (Loda *et al*. 2020, Sánchez-Ledesma *et al*. 2020), and commercial gentrification in New York’s Little Italy neighbourhoods, where retail change occurs but is disconnected from residential gentrification (Kosta 2019).

All studies described changes in the orientation of food businesses, finding that the retail food environment transformed to meet the needs of tourists or commuting workers at the expense of stores serving the everyday needs of residents.

None of the three studies looked at other aspects of the food environment, however since food outlets may adapt to tourist palates at the expense of locally preferred options, the issue of cultural preferences may be relevant.

## 7 Limitations

This REA has several limitations. *Food environment* is a relatively new term (Campeau *et al*. 2019), therefore relevant publications exploring concepts such as affordability or convenience, but not using *food environment* or other selected search terms, may have been missed.

While the concept of affordability was interpreted subjectively, with the inclusion of one study (Whittle et al 2015) showing how increased cost of living impacted affordability of food through purchasing power, the search terms used did not explicitly seek articles investigating the link between gentrification and cost of living. Therefore, studies highlighting this pathway will likely have been missed.

Only articles in English were included, however Morrison *et al*. (2009) found that limiting searches to English publications risks producing biased results. Since Western Europe was the second most represented geographic area, other relevant studies published in European languages could have been missed. The exclusion of articles in Spanish will likely have missed relevant studies from South American countries where urbanization and gentrification in the context of nutrition transitions are a concern.

Finally, concessions and adaptations made to the DFID *How To Note*, such as removing ‘reliability’ from the checklist, could have introduced bias, and the absence of alternative spelling of search terms was also a limitation of the search strategy. This assessment will also be prone to the usual selection bias of REAs due to the compromises required for them to be carried out rapidly (Barends *et al*. 2017).

## 8 Conclusion

This REA explored the question: *How does gentrification impact the healthfulness of food environments?* Through assessment of ten peer-reviewed studies, it found limited evidence that gentrification is associated with unhealthier food environments. The evidence body is small, comprised mostly of low to medium quality observational studies, albeit with consistent findings.

The exclusive use of observational study designs was considered appropriate for the research questions, but several limitations were identified nonetheless, including issues with measuring both gentrification and food environments, the classification of outlets broadly as ‘healthy’ or ‘unhealthy’, the use of cross-sectional data to answer a cause-and-effect research question, and inadequate control of confounding.

Of the four domains of food environments – availability, affordability, promotion, and food safety/quality/desirability – the first two were the most represented.

Past research such as James et al. (2017) has highlighted that whilst cross-sectionally, high-income neighbourhoods tend to have healthier food environments than low-income neighbourhoods, high-income neighbourhoods have become more unhealthy over time, whereas low-income neighbourhoods have plateaued. The results of this review add to this literature, finding that originally low-income neighbourhoods may mirror longitudinal trends of high-income neighbourhoods, developing more unhealthy food environments over time as they gentrify.

Downs *et al*. (2020)’s conceptual framework proposes that food environments transition with development, and that those in high-income developed urban societies may undergo further transition as consumers begin to demand healthy and sustainable foods. Viewing the current findings through this framework could imply that while gentrifying neighbourhoods may be undergoing this transition objectively, low income residents may simultaneously be experiencing a shift to unhealthier personal food environments.

The literature on affordability adds an important element to the food desert discourse, with food mirages behaving as food deserts in practical terms. However, affordability studies did not differentiate between healthy and unhealthy foods.

The theme of cultural acceptability (desirability) emerged, highlighting a gap in both the research and current conceptualization of food environments (Caspi et al. 2012). The impact of transient populations on food environments also arose, but further study into the impact on the cultural acceptability of foods would be relevant. The dimension of promotion did not feature at all in the research, nor did other food environment concepts such as quality, safety and convenience.

The geographical bias towards North America and Western Europe is representative of gentrification literature in general (Krase and DeSena 2020). However, given the increasing globalization of gentrification (Tulier et al. 2019), research in different regions could help isolate the causal effect of gentrification and control for locally contextual confounding factors.

Given the limitations presented in the REA, there remains significant room for improvement in research on gentrification and food environments. However, limited evidence should not be an excuse for inaction: urban policies that ensure the availability of healthy, affordable and culturally appropriate food should be pursued regardless, and are in line with every country’s commitment to Sustainable Development Goals 2 (zero hunger) and 11 (sustainable cities and communities).

Simultaneously, improvement in the evidence base can help policymakers better understand drivers of urban health inequalities and inform effective targeting of actions to achieve these goals.

The authors declared that they have no conflict of interest.

## Data Availability

All data produced in the present study are available upon reasonable request to the authors

## Statements and Declarations

We have no competing interests to declare.

